# Sleep Quality mediates the association between Tea Consumption and Duration of COVID-19-related Symptoms in the Elderly

**DOI:** 10.1101/2024.06.06.24308573

**Authors:** Yuxing Fan, Yunyu Wang, Jun Jiang, Shaopeng Yang, Jie Lu, Qinghua Ma, Hong Zhu

**Affiliations:** School of Public Health, Medical College of Soochow University, Suzhou 215123, China; The 3rd People’ s Hospital of Xiangcheng District, Suzhou 215134, China; School of Public Health, North China University of Science and Technology, Tangshan 063210, China; Anhui Medical College, Hefei 230601, China; Binzhou Zhanhua District Center for Disease Control and Prevention, Binzhou 256800, China

## Abstract

The association between tea consumption and the duration of COVID-19-related symptoms remains inconclusive. This cross-sectional study aims to investigate the potential mediating role of sleep quality in this association.

**Methods:** We conducted a cross-sectional study using data from elderly individuals aged 50 and above in Weitang Town in 2023. Detailed information on tea consumption, duration of COVID-19-related symptoms, and sleep quality was collected using pre-designed questionnaires. Sleep quality was assessed using the Pittsburgh Sleep Quality Index (PSQI). Spearman correlation analysis was employed to examine the relationships between variables. Mediation effect analysis utilized a mediation model with multi-category independent variables.

**Results:** The correlation analysis revealed negative associations between tea drinking frequency, type, years, concentration, and sleep quality. Additionally, sleep quality was significantly and positively associated with the duration of COVID-19-related symptoms, whereas various tea consumption variables were significantly and negatively associated with the duration of COVID-19-related symptoms. The mediation analysis confirmed that sleep quality partially mediated the relationship between daily tea drinking and the duration of COVID-19-related symptoms. Moreover, sleep quality fully mediated the associations between drinking green tea, consuming tea for less than 15 years or more than 30 years, the concentration of tea consumption, and the duration of COVID-19-related symptoms.

**Conclusions:** Tea consumption indirectly influences the duration of COVID-19-related symptoms through its impact on sleep quality. These findings highlight the importance of considering the effects of tea consumption on COVID-19 infection, as well as the potential to reduce the duration of COVID-19-related symptoms by improving sleep quality.

## 1. Introduction

The COVID-19 pandemic has presented an unprecedented public health and hygiene crisis. According to the World Health Organization, as of November 2023, there have been over 770 million reported cases of COVID-19 worldwide[1]. While most infected individuals experience mild symptoms[2], some symptoms can persist for an extended period[3]. Common symptoms include fever, nasal congestion, sore throat, hoarse voice, muscle pain, headache, among others. Research has indicated that long-term clinical manifestations following the acute phase of the illness can result in chronic inflammation and immune dysregulation[4,5].

Tea is widely enjoyed and has gained considerable attention. As a natural food, tea has been utilized in ancient China for treating certain infectious diseases. In modern times, drinking tea offers numerous health benefits, including the prevention of type 2 diabetes, obesity, cardiovascular disease, cancer, and immune-related diseases. These benefits are attributed to tea’s anti-inflammatory, immunomodulatory, and antioxidant properties[6,7] Additionally, tea can promote sleep by inducing a calming effect on the nerves[8]. Research has also demonstrated that tea polyphenols can regulate intestinal flora, potentially playing a role in preventing COVID-19[9]. Quality sleep is crucial for maintaining a robust immune system, as poor sleep quality increases susceptibility to viral infections[10]. Studies have indicated that COVID-19 patients with inadequate sleep experience prolonged duration of symptoms[11].

Both tea consumption and sleep quality have an impact on the duration of COVID-19-related symptoms, and they mutually influence each other. However, the underlying mechanism remains unclear. In our study, we hypothesized that individuals who consume tea would experience a reduced duration of COVID-19-related symptoms, with sleep quality playing a mediating role. Therefore, this project aims to investigate the relationship between tea consumption, sleep quality, and the duration of COVID-19-related symptoms.

## 2. Materials and methods

### 2.1 Study subjects

The study included elderly individuals aged 50-90 years old from Weitang Town, Suzhou City. Exclusion criteria were applied to those who were unable to provide the necessary information for the study or had incomplete information. A total of 2395 older adults were included in the study. All researchers agreed to administer the questionnaire survey and obtained signed informed consent forms. Ethical approval was obtained from the Research Ethics Committee of the Third People’s Hospital of Xiangcheng District, Suzhou City (Data protection number: xcsyllpj2023-001).

### 2.2 Measures and variables

#### 2.2.1 Basic demographic characteristics

Demographic information includes: age, sex, height, body weight, Body mass index (BMI).

#### 2.2.2 Tea consumption

Tea consumption was assessed based on the frequency, type, duration, and concentration of tea intake. Frequency of tea consumption was categorized as daily (≥1 cup/day), occasional (≥1 cup/week or ≥1 cup/month), rarely (<1 cup/month), or never. The types of tea consumed were classified as green tea or non-green tea. Duration of tea consumption was categorized as <15 years, 15-30 years, and ≥30 years. Tea concentration was determined by the proportion of tea leaves in the cup after brewing, with <25% considered light tea, 25% to 50% considered medium strength tea, and >50% considered strong tea[12].

#### 2.2.3 Sleep quality

The Pittsburgh Sleep Quality Index (PSQI) is employed to evaluate sleep quality, consisting of seven components: subjective sleep quality, sleep latency, sleep duration, habitual sleep efficiency, sleep disturbances, use of sleep medication, and daytime dysfunction. Total PSQI scores were calculated by summing the scores of these seven components. Scores can range from 0 to 21, with higher scores indicating poorer sleep quality[13].

#### 2.2.4 Duration (days) of COVID-19-related symptoms

The duration (in days) of COVID-19-related symptoms refers to the period during the pandemic when symptoms occur following infection with COVID-19. These symptoms encompass fever, nasal congestion, sore throat, hoarse voice, muscle pain, headache, cough, difficulty breathing, heart palpitations, vomiting, skin rashes, changes in vision, altered smell, loss of smell, and diarrhea.

### 2.3 Statistical analyses

The data were analyzed using IBM SPSS Statistics 27.0. Normally distributed continuous variables are presented as mean ± standard deviation, non-normally distributed continuous variables are reported as median (interquartile range), and categorical variables are presented as counts and percentages [n (%)]. Differences between males and females were assessed using t-tests, chi-square tests, and rank sum tests. The Spearman correlation method was employed to examine the relationships between tea consumption frequency, type, duration, concentration, sleep quality, and duration of COVID-19-related symptoms. Mediation analysis for multi-class independent variables was conducted using Model 4 of PROCESS, developed by Hayes, with Bootstrapping[14]. All tests were two-tailed, and statistical significance was set at *p* ≤ 0.05.

## 3. Results

### 3.1 Basic information about the research object

The participants had an average age of 67.68±0.29 years, with men comprising 41.2% of the sample. The participants had a mean BMI of 23.57±0.06 kg/m^2^. The average sleep quality score was 4.34±0.07. The median duration of COVID-19-related symptoms was 4.00 (1.00, 7.00) days. Male participants were significantly older than female participants (males: 70.32±0.17 years; females: 65.82±0.21 years; *p*<0.05). There were no significant differences, except for BMI, between male and female subgroups. Significant differences in tea consumption, sleep quality, and duration of COVID-19-related symptoms were observed between these two subgroups (Table 1).

**Table 1.**
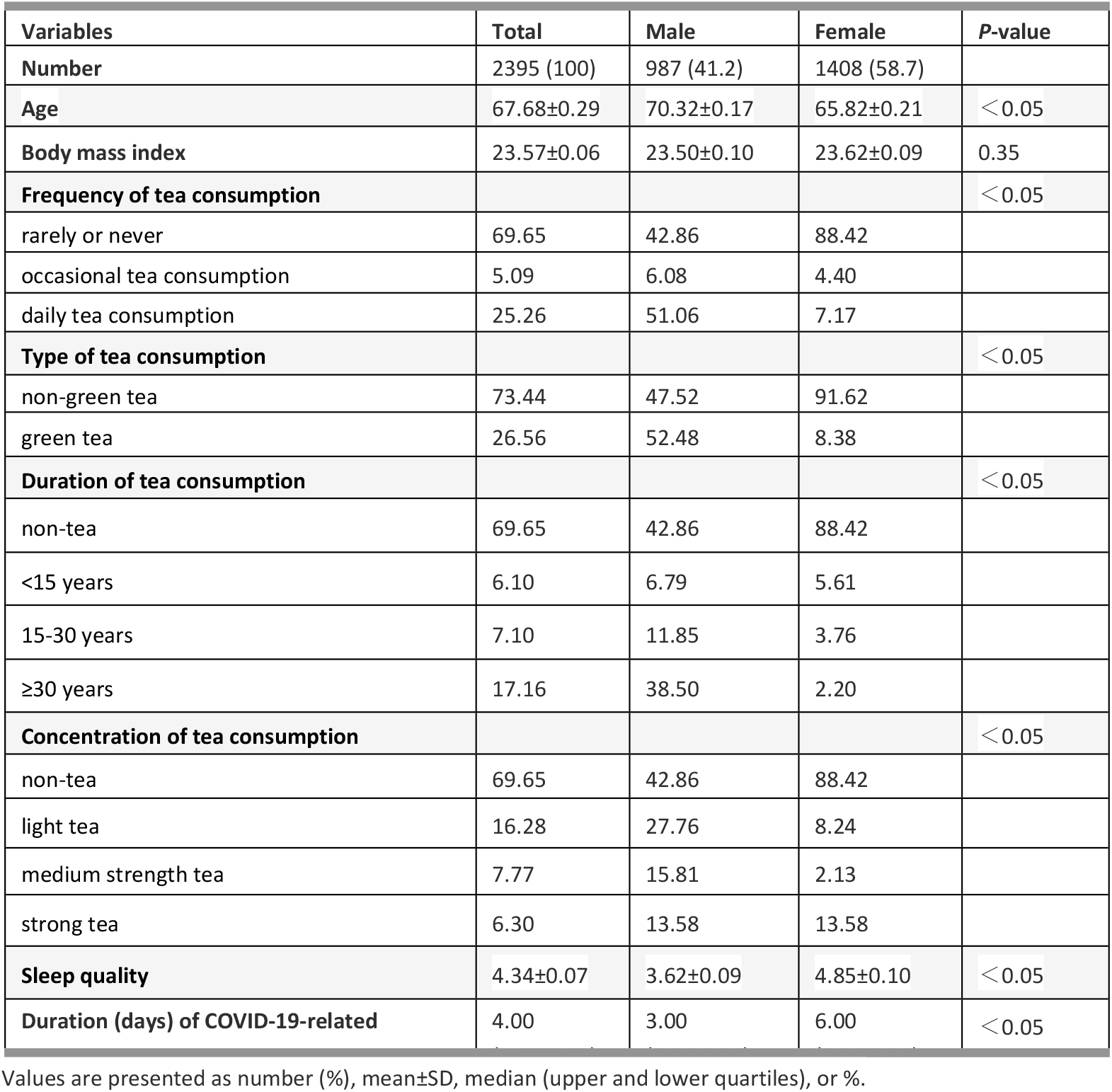
Basic information about the research object.

| Variables | Total | Male | Female | P-value |
| --- | --- | --- | --- | --- |
| **Number** | 2395 (100) | 987 (41.2) | 1408 (58.7) |  |
| **Age** | $67.68 \pm 0.29$ | $70.32 \pm 0.17$ | $65.82 \pm 0.21$ | $< 0.05$ |
| **Body mass index** | $23.57 \pm 0.06$ | $23.50 \pm 0.10$ | $23.62 \pm 0.09$ | 0.35 |
| **Frequency of tea consumption** | | | | $< 0.05$ |
| rarely or never | 69.65 | 42.86 | 88.42 |  |
| occasional tea consumption | 5.09 | 6.08 | 4.40 |  |
| daily tea consumption | 25.26 | 51.06 | 7.17 |  |
| **Type of tea consumption** | | | | $< 0.05$ |
| non-green tea | 73.44 | 47.52 | 91.62 |  |
| green tea | 26.56 | 52.48 | 8.38 |  |
| **Duration of tea consumption** | | | | $< 0.05$ |
| non-tea | 69.65 | 42.86 | 88.42 |  |
| <15 years | 6.10 | 6.79 | 5.61 |  |
| 15-30 years | 7.10 | 11.85 | 3.76 |  |
| $\geq 30$ years | 17.16 | 38.50 | 2.20 | |
| **Concentration of tea consumption** | | | | $< 0.05$ |
| non-tea | 69.65 | 42.86 | 88.42 |  |
| light tea | 16.28 | 27.76 | 8.24 |  |
| medium strength tea | 7.77 | 15.81 | 2.13 |  |
| strong tea | 6.30 | 13.58 | 13.58 |  |
| **Sleep quality** | $4.34 \pm 0.07$ | $3.62 \pm 0.09$ | $4.85 \pm 0.10$ | $< 0.05$ |
| **Duration (days) of COVID-19-related** | 4.00 | 3.00 | 6.00 | $< 0.05$ |

Values are presented as number (%), mean ± SD, median (upper and lower quartiles), or %.

### 3.2 Correlation analysis

A Spearman correlation analysis was conducted to examine the relationship between tea consumption, sleep quality, and duration of COVID-19-related symptoms (Table 2). The results indicated that tea consumption frequency, type, duration, and concentration were negatively correlated with sleep quality (*r*=-0.107, *r*=-0.105, *r*=-0.102, *r*=-0.098; all *p*-values < 0.01). This suggests that higher tea consumption frequency, consumption of green tea, longer duration of tea consumption, and higher concentration of tea consumption are associated with better sleep quality. Additionally, sleep quality was positively correlated with the duration of COVID-19-related symptoms (*r*=0.204, *p*<0.001), indicating that poor sleep quality is linked to a longer duration of COVID-19-related symptoms. Moreover, there was a negative correlation between tea consumption frequency, type, duration, concentration, and the duration of COVID-19-related symptoms (*r*=-0.145, *r*=-0.145, *r*=-0.150, *r*=-0.144; all *p*-values < 0.01). In other words, higher tea consumption frequency, consumption of green tea, longer duration of tea consumption, and higher concentration of tea consumption were associated with a shorter duration of COVID-19-related symptoms.

**Table 2.**
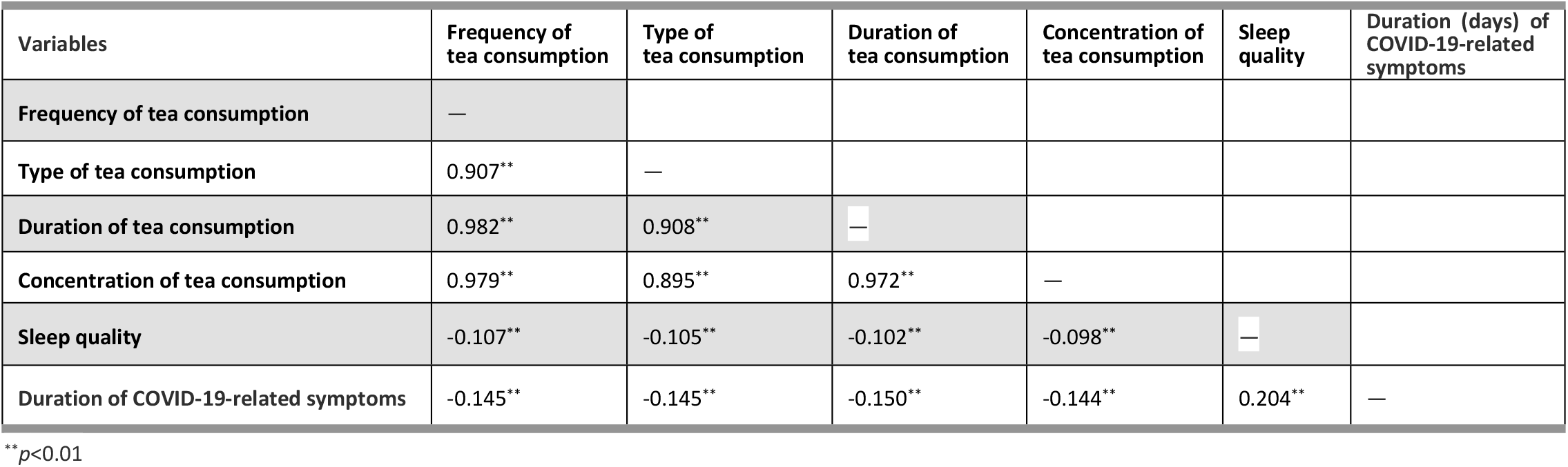
Spearman correlation analysis among perceived tea consumption, sleep quality, and duration of COVID-19-related symptoms.

| Variables | Frequency of tea consumption | Type of tea consumption | Duration of tea consumption | Concentration of tea consumption | Sleep quality | Duration (days) of COVID-19-related symptoms |
| --- | --- | --- | --- | --- | --- | --- |
| Frequency of tea consumption | — |  |  |  |  |  |
| Type of tea consumption | 0.907** | — |  |  |  |  |
| Duration of tea consumption | 0.982** | 0.908** | — |  |  |  |
| Concentration of tea consumption | 0.979** | 0.895** | 0.972** | — |  |  |
| Sleep quality | -0.107** | -0.105** | -0.102** | -0.098** | — |  |
| Duration of COVID-19-related symptoms | -0.145** | -0.145** | -0.150** | -0.144** | 0.204** | — |

\*\* $p<0.01$

### 3.3 Results of mediation analysis

#### 3.3.1 The mediating role of sleep quality between frequency of tea consumption and duration of COVID-19-related symptoms

Relative mediation analysis revealed a significant relative mediation effect when comparing daily tea drinkers to non-tea drinkers, with a 95% confidence interval of [-0.7478, -0.2343]. This indicates that daily tea consumption is associated with higher sleep quality and a reduced duration of COVID-19-related symptoms (*a*_*2*_=-0.9076, *b*=0.5084, *a*_*2*_*b*=-0.4614). Moreover, the relative direct effect was significant (*c*_*2*_*’*=-2.1268, *p*<0.001), indicating that daily tea drinkers have a shorter duration of COVID-19-related symptoms compared to non-tea drinkers when the mediating effect is excluded. The relative total effect was also significant (*c*_*2*_=*a*_*2*_*b*+*c*_*2*_*’*=-2.5882, *p*<0.001), with the relative mediation effect accounting for 17.8% of the total effect size (0.4614/2.5882). Conversely, when comparing occasional tea drinkers to non-tea drinkers, the 95% confidence interval was [-0.3724, 0.3053], indicating no significant relative mediation effect (*a*_*1*_=-0.0819, *b*=0.5084, *a*_*1*_*b*=-0.0417). For detailed information, refer to Figure 1 and Table 3.

**Table 3.** Multicategory mediation modeling: mediating effects of sleep quality in the influence of occasional and daily tea consumption on the duration of COVID-19-related symptoms, n=2395.

| Trails | Unstandardized coefficients(S.E.) | P | 95% confidence interval |
| --- | --- | --- | --- |
| **sleep quality** |  |  |  |
| Occasional tea consumption ($a_1$) | -0.0819(0.3170) | 0.7961 | -0.7036,0.5397 |
| Daily tea consumption ($a_2$) | -0.9076(0.1604) | <0.001 | -1.2222,-0.5931 |
| **Duration of COVID-19-related** |  |  |  |
| sleep quality ($b$) | 0.5084(0.1152) | <0.001 | 0.2825,0.7342 |
| Occasional tea consumption ($c_1'$) | 2.0358(1.7860) | 0.2545 | -1.4665,5.5380 |
| Daily tea consumption ($c_2'$) | -2.1268(0.9098) | 0.0195 | -3.9108,-0.3427 |
| **Relative indirect effects** |  |  |  |
| $a_1b$ | -0.0417(0.1660) | — | -0.3724,0.3053 |
| $a_2b$ | -0.4614(0.1302) | — | -0.7478,-0.2343 |
| **Relative total effects** |  |  |  |
| $a_1b+c_1'$ | 1.9941(1.7929) | 0.2661 | -1.5216,5.5098 |
| $a_2b+c_2'$ | -2.5882(0.9072) | 0.0044 | -4.3672,-0.8091 |

**Figure 1.**
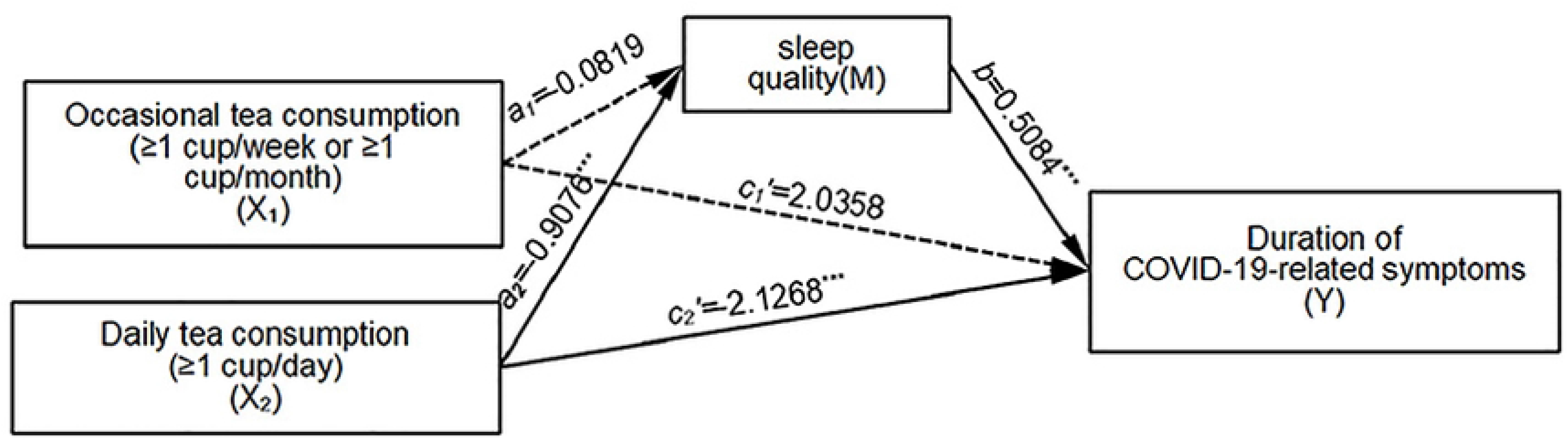
Sleep quality serves as a mediator between tea drinking frequency and the duration of COVID-19-related symptoms. Unstandardized coefficients are presented. The solid line represents a significant mediating effect, while the dotted line indicates a non-significant mediating effect. ^***^*p*<0.001.

#### 3.3.2 The mediating role of sleep quality between type of tea consumption and duration of COVID-19-related symptoms

We utilized Model 4 in PROCESS to examine the impact of sleep quality on the relationship between tea consumption type and the duration of COVID-19-related symptoms. The findings revealed that the direct prediction of the duration of COVID-19-related symptoms by tea drinking type was not significant (*c’*=-1.6025, *p*=0.0709). When comparing green tea drinkers to non-drinkers, with non-drinking of green tea as the reference level, the 95% confidence interval was [-0.7071, -0.2181], indicating a significant mediation effect (*a*=-0.8358, *b*=0.5184, *ab*=-0.4333). This implies that individuals who consume green tea have higher sleep quality and experience a reduced duration of COVID-19-related symptoms. The direct effects were found to be non-significant, suggesting complete mediation. Moreover, the total effect was significant (*c*=*ab*+*c’*=-2.0358, *p*<0.001), with the mediation effect accounting for 21.3% of the total effect size (0.4333/2.0358). For detailed information, refer to Figure 2.

**Figure 2.**
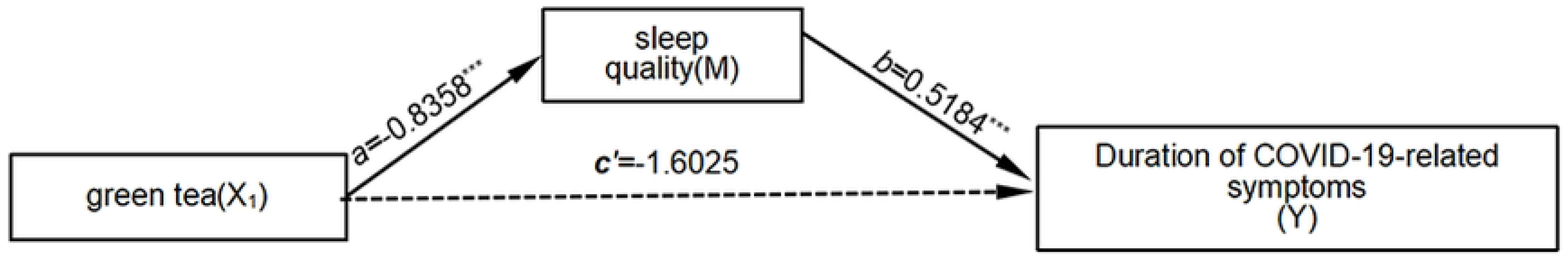
Sleep quality acts as a mediator between the type of tea consumption and the duration of COVID-19-related symptoms. Unstandardized coefficients are provided. The solid line signifies a significant mediating effect, while the dotted line indicates a non-significant mediating effect. ^***^*p*<0.001.

#### 3.3.3 The mediating role of sleep quality between tea consumption duration and COVID-19-related symptoms

Relative mediation analysis revealed a significant mediating effect when comparing tea drinking for less than 15 years and tea drinking for more than 30 years to non-tea drinking (*a*_*1*_=-0.6628, *b*=0.5188, *a*_*1*_*b*=-0.3439; *a*_*3*_=-0.8973, *b*=0.5188, *a*_*3*_*b*=-0.4656). This indicates that individuals who have been consuming tea for less than 15 years and those who have been consuming tea for more than 30 years exhibit better sleep quality compared to non-tea drinkers. Moreover, both groups experience a corresponding reduction in COVID-19-related symptoms. The relative direct effects are not significant, suggesting a complete mediation effect. The relative total effect of consuming tea for less than 15 years, compared to non-tea drinking, is not significant. However, the relative total effect of individuals who have been consuming tea for more than 30 years, compared to non-tea drinking, is significant (*c*_*3*_=*a*_*3*_*b*+*c*_*3*_*’*=-2.5036, *p*<0.001). The effect size of the relative mediation effect is 18.6% (0.4656/2.5036). It is important to note that sleep quality does not mediate the relationship between tea drinking for 15-30 years and COVID-19-related symptoms. For further details, refer to Figure 3.

**Figure 3.**
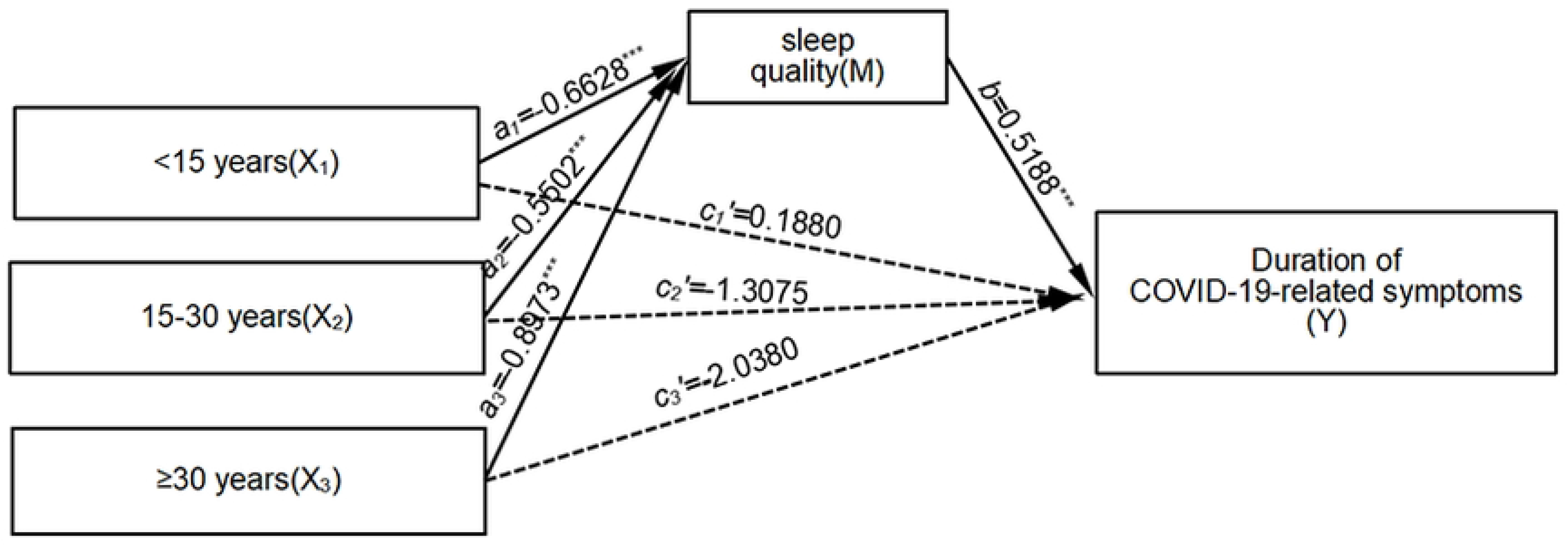
The duration of COVID-19-related symptoms is mediated by sleep quality. Unstandardized coefficients are provided. The solid line represents a significant mediating effect, while a dotted line represents a non-significant mediating effect. ^***^*p*<0.001.

#### 3.3.4 The mediating role of sleep quality between concentration of tea consumption and duration of COVID-19-related symptoms

Relative mediation analysis revealed a significant mediating effect when comparing drinking light tea, moderate tea, and strong tea to not drinking tea, using non-tea drinking as the reference level (*a*_*1*_=-0.6943, *b*=0.5218, *a*_*1*_*b*=-0.3623; *a*_*2*_=-0.9555, *b*=0.5218, *a*_*2*_*b*=-0.4986; *a*_*3*_=-0.7327, *b*=0.5188, *a*_*3*_*b*=-0.3823). This indicates that individuals who consume light tea, moderate tea, and strong tea experience better sleep quality compared to non-tea drinkers. Additionally, they also exhibit a reduction in COVID-19-related symptoms. The relative direct effects are not significant, suggesting a complete mediation effect. The relative total effect of drinking light tea and moderate tea, compared to not drinking tea, is not significant. However, the relative total effect of drinking strong tea, compared to not drinking tea, is significant (*c*_*3*_=*a*_*3*_*b*+*c*_*3*_*’*=-3.3876, *p*<0.001). The effect size of the relative mediation effect is 11.3% (0.3823/3.3876). For further details, refer to Figure 4.

**Figure 4.**
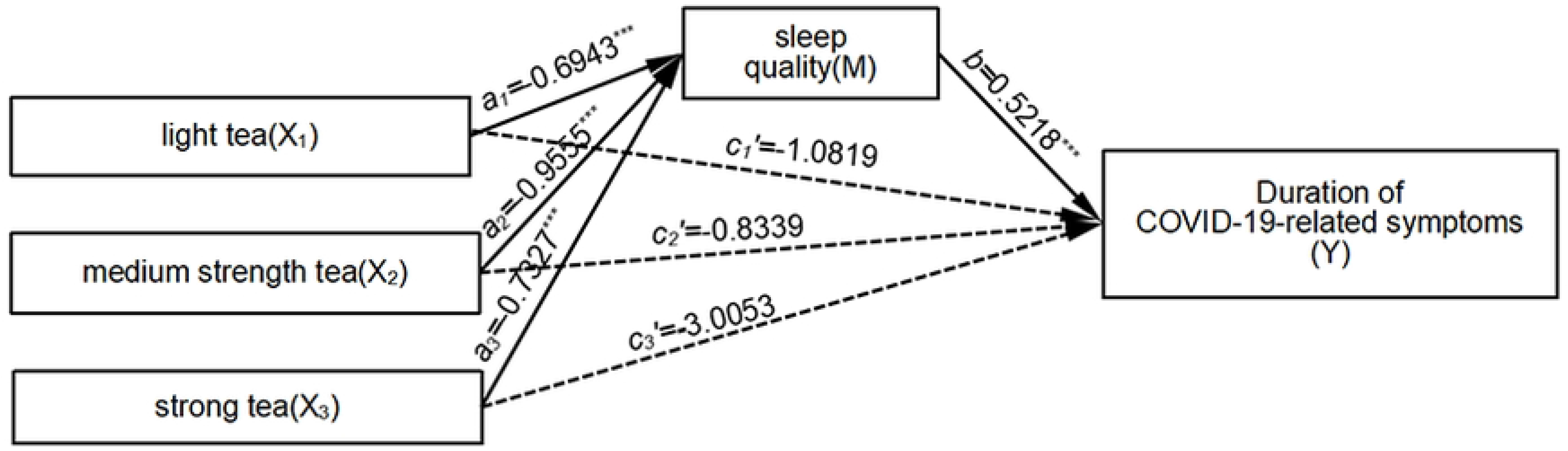
Sleep quality acts as a mediator between the concentration of tea consumption and the duration of COVID-19-related symptoms. The unstandardized coefficients are provided, with a solid line indicating a significant mediating effect and a dotted line indicating a non-significant mediating effect. ^***^*p*<0.001.

## 4. Discussion

The COVID-19 pandemic has emerged as a significant public health crisis, impacting not only my country but also the global community. It has spread rapidly, affected a wide population, and posed challenges in terms of prevention and control efforts. This study retrospectively examined cases of Omicron variant infection during the December 2023 pandemic. The findings revealed that the median duration of COVID-19-related symptoms was 4.00 (1.00, 7.00) days, which was shorter compared to the median time of 10.3 (7.4, 12.4) days for nucleic acid tests to turn negative in a makeshift hospital in Shanghai in May 2022[15]. This difference can likely be attributed to the reduced virulence of the virus during the ongoing pandemic, resulting in less severe infections among individuals.

When examining tea consumption patterns, it becomes evident that a relatively small proportion of individuals, approximately 30.35%, consume tea. Among these tea consumers, the distribution between males and females is 57.14% and 11.58%, respectively. Furthermore, male tea drinkers exhibit significantly longer tea drinking durations and higher tea consumption concentrations, aligning with previous research findings[16]. The assessment of sleep quality revealed an average score of 4.34 points among participants, ranging from 0 to 5. This score indicates a higher sleep quality compared to the study conducted by Xiaolei Liu et al. in 2022 (PSQI score: 6.16±3.29)[17]. In essence, the overall sleep quality is deemed to be very good. These findings corroborate similar studies, consistently indicating that men tend to have significantly better sleep quality than women[18]. Consequently, interventions aimed at improving sleep quality should prioritize women and provide additional attention to this demographic.

In this study, we investigate the relationship between tea drinking and the duration of COVID-19-related symptoms in older adults. Additionally, we examine the mediating effect of sleep quality on this association. The findings indicate that sleep quality serves as a significant mediator between daily tea consumption, consumption of green tea, duration of tea drinking (less than 15 years and more than 30 years), concentration of tea consumption, and the duration of COVID-19-related symptoms.

Patients with COVID-19 commonly experience clinical symptoms, including gastrointestinal manifestations such as vomiting and diarrhea. Numerous investigations have demonstrated that COVID-19 patients with gastrointestinal symptoms exhibit a more rapid disease progression, increased likelihood of severe illness, and poorer survival outcomes compared to those without such symptoms[19]. Tea, a popular beverage rich in phytochemicals, is increasingly recognized for its potential health benefits and disease prevention properties, making it a favored choice for many individuals seeking a convenient and healthy lifestyle. Several studies have highlighted the ability of tea to regulate the gut microbiota and modulate both local and systemic immune responses in the intestines, potentially contributing to its role in mitigating COVID-19 to some extent[9]. These empirical findings align with the results of our study, which indicate that daily tea consumption is associated with a reduction in the duration of COVID-19-related symptoms.

Tea drinking behavior can impact sleep quality, thereby influencing COVID-19-related symptoms. This is attributed to the various health benefits of tea, such as promoting sleep, reducing stress, and regulating energy metabolism[20]. However, there are ongoing debates regarding the specific effects of tea type[8,21,22], frequency, and quantity on sleep quality. Among the different types of tea, green tea is particularly popular in Suzhou. Green tea is rich in bioactive compounds, including theanine, caffeine, and tea polyphenols. Theanine possesses anti-fatigue, sedative, and sleep-enhancing properties, counteracting the potential negative effects of caffeine on sleep quality[23]; Tea polyphenols act as antioxidants, effectively reducing inflammation and oxidative stress caused by sleep deprivation, thereby improving sleep quality and memory[24]. Tea polyphenols act as antioxidants, effectively reducing inflammation and oxidative stress caused by sleep deprivation, thereby improving sleep quality and memory[25]. Consistent with these findings, our study demonstrates that individuals who consume green tea exhibit better sleep quality. Furthermore, the duration of tea drinking also plays a significant role, with long-term tea consumption of 30 years or more positively impacting sleep quality. Habitual tea drinking is associated with an improved health-related quality of life in older Chinese adults, which aligns with our data findings on the relationship between tea drinking and sleep quality[26]. Moreover, research suggests that the benefits of tea may be amplified by consuming stronger tea varieties with higher concentrations of beneficial compounds[27].

The duration of COVID-19 symptoms is inversely related to sleep quality. A prospective association exists between sleep disturbance and the development of long-term symptoms after COVID-19. Lower sleep quality, more severe insomnia, and shorter sleep duration 1 to 3 months post-COVID-19 are associated with higher odds of developing a wide range of clinical manifestations. Our findings suggest that the impact of sleep loss on vaccination effectiveness may mediate the link between sleep and post-COVID-19 symptoms[11]. Moreover, studies have reported the efficacy of various tea ingredients in blocking COVID-19 infection[28]. Tea polyphenols, for example, can inhibit viruses by positively influencing the gut microbiota. Additionally, increased tea consumption has been associated with a reduced risk of COVID-19 infection[6]. Based on the aforementioned research, it can be inferred that enhancing tea drinking behavior can improve sleep quality and consequently mitigate the impact of COVID-19 symptoms on the human body. Therefore, tea consumption not only affects post-COVID-19 symptoms by regulating the internal intestinal flora, as observed in previous studies, but also indirectly influences these symptoms through its impact on sleep.

This study has several limitations. Firstly, due to the nature of the cross-sectional design, our study was unable to establish a causal relationship between tea consumption, sleep quality, and duration of COVID-19 symptoms. Future research could employ a longitudinal design to address this limitation. Secondly, the information regarding tea drinking, sleep, and COVID-19 was collected through self-reporting methods, which may be subject to retrospective inaccuracies. In future studies, objective measurements such as sleep monitoring watches could be utilized to assess sleep quality. Thirdly, our research subjects were limited to residents of Weitang Town, which exhibits a certain geographical and population concentration. Therefore, a larger sample size may be necessary to generalize the findings to the entire country.

Our study indicates that sleep quality plays a partial mediating role in the relationship between tea consumption and COVID-19-related symptoms among the elderly. As a result, our findings not only contribute to a deeper understanding of the mechanism by which tea consumption affects the duration of COVID-19-related symptoms, but also offer new insights for the prevention and treatment of COVID-19 infection.

## Data Availability

Data cannot be shared publicly because of participants did not provide their consent to share the data.

## Author Contributions

**Conceptualization:** Yuxing Fan, Qinghua Ma.

**Data curation:** Yuxing Fan, Yunyu Wang, Jun Jiang.

**Formal analysis:** Yuxing Fan.

**Supervision:** Shaopeng Yang, Jie Lu,Qinghua Ma,Hong Zhu.

**Writing – original draft:** Yuxing Fan, Jun Jiang.

**Writing – review & editing:** Yuxing Fan, Shaopeng Yang, Jie Lu,Qinghua Ma,Hong Zhu.

## References

1. Organization W H 2023 Who coronavirus (covid-19) dashboard https://covid19.who.int/.

2. Gao C, Feng F J, Jiang J J, Yao X W, Yu X H and Zhang J C 2022 The latest research progress of the corona virus disease 2019 mutant strain “omicron” Journal of Hainan Medical University 28(7) 481–5

3. Davis H E, Assaf G S, Mccorkell L, Wei H, Low R J, Re’Em Y, Redfield S, Austin J P and Akrami A 2021 Characterizing long covid in an international cohort: 7 months of symptoms and their impact Eclinicalmedicine 38 101019

4. Mazza M G, Palladini M, De Lorenzo R, Magnaghi C, Poletti S, Furlan R, Ciceri F, Rovere-Querini P and Benedetti F 2021 Persistent psychopathology and neurocognitive impairment in covid-19 survivors: effect of inflammatory biomarkers at three-month follow-up Brain Behav Immun 94 138–47

5. Phetsouphanh C et al 2022 Immunological dysfunction persists for 8 months following initial mild-to-moderate SARS-cov-2 infection Nat Immunol 23(2) 210–6

6. Baranova A, Song Y, Cao H, Yue W and Zhang F 2022 Causal associations of tea intake with covid-19 infection and severity Front. Nutr. 9 1005466

7. Tang G Y et al. 2019 Health functions and related molecular mechanisms of tea components: an update review Int J. Mol Sci 20(24) 6196

8. Ouyang J, Peng Y and Gong Y 2022 New perspectives on sleep regulation by tea: harmonizing pathological sleep and energy balance under stress Foods 11(23) 3930

9. Xiang Q, Cheng L, Zhang R, Liu Y, Wu Z and Zhang X 2022 Tea polyphenols prevent and intervene in covid-19 through intestinal microbiota Foods 11(4) 506

10. Besedovsky L, Lange T and Born J 2012 Sleep and immune function Pflugers Arch 463(1) 121–37

11. Salfi F, Amicucci G, Corigliano D, Viselli L, D’Atri A, Tempesta D and Ferrara M 2023 Poor sleep quality, insomnia, and short sleep duration before infection predict long-term symptoms after covid-19 Brain Behav Immun 112 140–51

12. Jie J L, Tong J H, Huang L P, He F, Xu Q P, Xiong W M and Cai L 2019 Association of tea and dairy products consumption with primary lung cancer among people neither smoking nor drinking alcohol: a case-control study Chin J Public Health 35(11) 1496–500

13. Zerón-Rugerio M F, Hernáez Á, Cambras T and Izquierdo-Pulido M 2022 Emotional eating and cognitive restraint mediate the association between sleep quality and bmi in young adults Appetite 170 105899

14. Fang J, Wen Z L and Zhang M Q 2017 Mediation analysis of categorical variables Journal of Psychological Science 40(2) 471–7

15. Chen Y L, Chen Z, Wang X H, Xiong H, Shuang F and Liu X J 2022 Influencing factors of nucleic acid negative conversion in patients with mild and common covid-19 induced by the omicron variant of SARS-cov-2 Journal of Zhejiang University(Medical Sciences) 51(6) 731–7

16. Tian T et al 2020 Tea consumption and risk of stroke in chinese adults: a prospective cohort study of 0.5 million men and women Am J. Clin Nutr 111(1) 197–206

17. Liu X, Xia X, Hu F, Hao Q, Hou L, Sun X, Zhang G, Yue J and Dong B 2022 The mediation role of sleep quality in the relationship between cognitive decline and depression Bmc Geriatr. 22(1) 178

18. Hu W et al 2022 The role of depression and physical activity in the association of between sleep quality, and duration with and health-related quality of life among the elderly: a uk biobank cross-sectional study Bmc Geriatr. 22(1) 338

19. Zhu H, Wang L, Fang C, Peng S, Zhang L, Chang G, Xia S and Zhou W 2020 Clinical analysis of 10 neonates born to mothers with 2019-ncov pneumonia Transl. Pediatr. 9(1) 51–60

20. Hou S J, Tsai S J, Kuo P H, Lin W Y, Liu Y L, Yang A C, Lin E and Lan T H 2021 An association study in the taiwan biobank elicits the gabaa receptor genes gabrb3, gabra5, and gabrg3 as candidate loci for sleep duration in the taiwanese population Bmc Med. Genomics 14(1) 223

21. Hou S J, Tsai S J, Kuo P H, Liu Y L, Yang A C, Lin E and Lan T H 2020 An association study in the taiwan biobank reveals rora as a novel locus for sleep duration in the taiwanese population Sleep Med 73 70–5

22. Althakafi K A, Alrashed A A, Aljammaz K I, Abdulwahab I J, Hamza R, Hamad A F and Alhejaili K S 2019 Prevalence of short sleep duration and effect of co-morbid medical conditions - a cross-sectional study in saudi arabia J. Family Med. Prim. Care 8(10) 3334–9

23. Baba Y, Takihara T and Okamura N 2023 Theanine maintains sleep quality in healthy young women by suppressing the increase in caffeine-induced wakefulness after sleep onset Food Funct. 14(15) 7109–16

24. Romain C, Alcaraz P E, Chung L H and Cases J 2017 Regular consumption of holisfiit, a polyphenol-rich extract-based food supplement, improves mind and body well-being of overweight and slightly obese volunteers: a randomized, double-blind, parallel trial Int J. Food Sci Nutr 68(7) 840–8

25. Li X et al 2017 Tea consumption and risk of ischaemic heart disease Heart 103(10) 783–9

26. Vieux F, Maillot M, Rehm C D and Drewnowski A 2019 Tea consumption patterns in relation to diet quality among children and adults in the united states: analyses of nhanes 2011-2016 data Nutrients 11(11) 2635

27. Li F D, He F, Ye X J, Shen W, Wu Y P, Zhai Y J, Wang X Y and Lin J F 2016 Tea consumption is inversely associated with depressive symptoms in the elderly: a cross-sectional study in eastern china J. Affect Disord 199 157–62

28. Liu J et al 2021 Epigallocatechin gallate from green tea effectively blocks infection of SARS-cov-2 and new variants by inhibiting spike binding to ace2 receptor Cell Biosci. 11(1) 168

